# Diabetes and Incidence of Breast Cancer and its Molecular Subtypes: A Systematic Review and Meta-Analysis

**DOI:** 10.1101/2023.05.13.23289893

**Authors:** Fanxiu Xiong, Qichen Dai, Sihan Zhang, Stephen Bent, Peggy Tahir, Erin L. Van Blarigan, Stacey A. Kenfield, June M. Chan, Gabriela Schmajuk, Rebecca E. Graff

## Abstract

Diabetes mellitus (DM) has been proposed to be positively associated with breast cancer (BCa) risk due to shared risk factors, metabolic dysfunction, and use of antidiabetic medications. We conducted a systematic review and meta-analysis to evaluate the association between DM and BCa risk. We searched PubMed, Embase, and Web of Science for cohort and case-control studies assessing the association between DM and BCa published before December 10, 2021. Two reviewers independently screened the studies for inclusion, abstracted article data, and rated study quality. Random effects models were used to estimate summary risk ratios (RRs) and 95% confidence intervals (CIs). From 8396 articles identified in the initial search, 70 independent studies were included in the meta-analysis. DM was associated with an overall increased risk of BCa (RR=1.20, 95% CI: 1.11-1.29). The 24 case-control studies demonstrated a stronger association (RR=1.26, 95% CI: 1.13-1.40) than the 46 cohort studies (RR=1.15, 95% CI: 1.05-1.27). Studies reporting risk by menopausal status found that postmenopausal women had an elevated risk of developing BCa (RR=1.12, 95% CI: 1.07-1.17). No association between DM and BCa risk was observed among premenopausal women (RR=0.95, 95% CI: 0.85-1.05). In addition, DM was associated with significantly increased risks of estrogen receptor (ER)+ (RR=1.09, 95% CI: 1.00-1.20), ER- (RR=1.16, 95% CI: 1.04-1.30), and triple negative BCa (RR=1.41, 95% CI: 1.01-1.96). The association estimate for human epidermal growth factor 2- positive BCa was also positive (RR=1.21, 95% CI: 0.52-2.82), but the confidence interval was wide and crossed the null. Our meta-analysis confirms a modest positive association between DM and BCa risk. In addition, our results suggest that the DM and BCa association may be modified by menopausal status, and DM may be differentially associated with BCa subtypes defined by receptor status. Additional studies are warranted to investigate the mechanisms underlying these associations and any influence of DM on BCa receptor expression.

## 1 INTRODUCTION

Among women worldwide, breast cancer (BCa) is the most prevalent malignancy and the leading cause of cancer-related death[1]. In 2020, there were an estimated 2.3 million new cases of female BCa and more than half a million deaths from disease[2]. Obesity is an established risk factor for BCa, particularly in postmenopausal women[3]. Given the relationship between obesity and BCa, it stands to reason that varying metabolic abnormalities could be associated with BCa risk. Diabetes mellitus (DM) is one of the most common metabolic abnormalities globally, such that 463 million people were living with DM in 2019[4]. DM and its complications increase the risk of cardiovascular disease, neuropathy, retinopathy, kidney failure, and overall mortality[5], but its relationship with BCa has not been firmly established.

Due to shared risk factors, metabolic dysfunction in diabetic patients, and antidiabetic medication use, it has been proposed that DM could be positively associated with subsequent BCa risk[6]. Indeed, the potential role of DM in BCa incidence has been widely investigated. The largest existing meta-analysis, which was published in 2012 and included 40 observational studies, indicated a 20% excess risk of BCa among women with DM[7]. Heterogeneity among studies included in this meta-analysis was substantial, and studies published since have yielded inconsistent findings. In addition, studies investigating associations between DM and specific BCa molecular subtypes have been inconclusive. Given that BCa is a heterogeneous disease – often grouped according to joint expression of hormonal and growth receptors, including estrogen receptor (ER), progesterone receptor (PR), and human epidermal growth factor receptor 2 (HER2)[8] – it is important to consider possible distinctions in metabolic etiology.

A solidified understanding of the relationship between DM and the risk of BCa and its subtypes is important for clarifying BCa etiology and informing screening decisions. Therefore, we reviewed and synthesized the findings from relevant epidemiologic studies to achieve summary estimates for associations between DM and risk of BCa overall and of BCa defined by receptor expression.

## 2 MATERIALS AND METHODS

### 2.1 Research question

Toward the formulation of a coherent research question, we implemented the Population, Interventions or Exposure, Comparator, Outcome, Study Design (PICOS) approach, as suggested by PRISMA guidelines.

#### Population

Women aged 18 years or older.

#### Exposure

DM identified prior to any detection of BCa. Gestational DM, which only occurs during pregnancy, was not included.

#### Comparison

Lacking a diagnosis of DM prior to any detection of BCa.

#### Outcome

Malignant BCa, identified by self-report or from medical records.

#### Study Design

For our primary analyses, cohort and case-control studies were included. To assess associations between DM and BCa molecular subtypes, all other study designs were considered.

### 2.2 Literature search

This systematic review and meta-analysis was conducted according to the Preferred Reporting Item for Systematic Reviews and Meta-analysis 2020 (PRISMA 2020) statement[9]. The protocol was registered in the International Prospective Register of Systematic Reviews (PROSPERO CRD42022311936).

We conducted a comprehensive database search regarding DM and BCa risk in PubMed (National Library of Medicine), Embase (Elsevier), and Web of Science (Clarivate Analytics). We focused on three main concepts: diabetes, BCa, and associated risks and incidence. Appropriate synonyms were developed for each concept, and we used both keywords and index terms (e.g., Mesh, Emtree) appropriate to each database. Full search strategies are included in the Search Appendix. Human studies that were published by Dec 10, 2021 and written in English were included. We also reviewed the reference section of studies included in previously published systematic reviews regarding DM and BCa to identify additional studies that met the eligibility criteria[7,10–14].

### 2.3 Study selection

Titles and abstracts were independently screened by two reviewers (FX and QD) to identify potentially relevant epidemiologic studies. Full texts of potentially relevant articles were retrieved and reviewed according to inclusion (PICOS) and exclusion criteria (below). Disagreements that were not resolved by discussion between the two reviewers were adjudicated by a third reviewer (SZ).

Studies were excluded if: they did not differentiate between benign and malignant breast tumors; DM was diagnosed at the same time as or after BCa diagnosis; the number of BCa cases in cohort studies or DM cases in case-control studies was less than 10; they were not human studies; they were not written in English; or they were case reports, review articles, meeting abstracts, or letters. When more than one study was conducted in the same study population, only the most recent findings were included.

### 2.4 Data abstraction and quality assessment

Two reviewers (FX and SZ) independently abstracted data from relevant articles using a standardized form. Discrepancies were resolved by discussion with a third reviewer (QD). Abstracted information included: first author, year of publication, study population, study design, country of study, age range, follow-up length or calendar period, DM assessment method, DM classification (i.e., any DM, type 1 DM (T1D), and/or type 2 DM (T2D)), BCa assessment method, BCa molecular subtypes, covariates, risk estimates with corresponding 95% confidence intervals (CIs), and any association measures within subgroups. Only estimates from fully-adjusted models were abstracted.

The Newcastle-Ottawa-Scale (NOS) was employed to evaluate the risk of bias in selected cohort and case-control studies[15]. Briefly, the NOS consists of eight items and is categorized into three domains: 1) selection of study population (4 items); 2) level of comparability between the study groups (1 item); and 3) assessment of exposure for case-control studies or outcome for cohort studies (3 items). One star could be awarded for each item, with the exception of a possible two stars for the single comparability item. Studies with stars of 8-9, 6-7, and 1-5 were deemed high-, medium-, and low-quality, respectively.

### 2.5 Statistical analysis

If risk ratios (RRs) were not reported in contributing studies, we assumed that other risk measures, such as odds ratios (ORs) and hazard ratios (HRs), numerically approximated RRs under the rare disease assumption[16]. When contributing studies only reported subgroup estimates (e.g., separate estimates for pre- and post- menopausal women), we calculated summary estimates for these individual studies before including them in our meta-analysis. To do so, we used fixed effects models when heterogeneity between subgroups was not significant and random effects models when heterogeneity was significant (P < 0.05).

For our meta-analysis, we estimated summary RRs and 95% CIs using random effects models, to allow for differences in study populations and generate conservative results. Heterogeneity across studies was assessed using 1) the I^2^ statistic, with 0-25%, 25-75%, and >75% representing possibly unimportant, moderate, and substantial heterogeneity, respectively, and 2) Cochran’s Q statistic, with P < 0.05 as the threshold for statistical significance. Leave-one-out sensitivity analyses were carried out to evaluate the influence of each study on the overall summary estimate. A temporal cumulative meta-analysis was also performed to show how the overall estimate changed as each study was added in chronological order. We additionally performed analyses stratified by study quality (high, medium, or low).

Publication bias was graphically assessed using a funnel plot. Duval and Tweedie’s trim-and-fill method was implemented based on the funnel plot to estimate the influence of potentially missing studies on the summary estimate[17]. Begg’s test and Egger’s test were also implemented to assess publication bias.

To explore potential sources of heterogeneity, separate analyses were performed by study design (cohort study or case-control study), study region (Asia, Americas, Europe, or Australia), year of publication (before or after the largest existing review, published in 2012)[10], method of DM ascertainment (self-report or medical review), adjustment for body mass index (BMI; no or yes), and adjustment for parity (no or yes). Meta-regression analyses stratified by the aforementioned subgroups were then performed to examine differences.

Some studies performed separate analyses in pre- and post-menopausal women. We used random-effects models to estimate the association of DM with pre- and post-menopausal BCa separately. If no information regarding menopausal status was provided, we defined postmenopausal to be age 50 years or older[18]. In addition, we examined associations between DM and BCa molecular subtypes. The subtypes were grouped into four categories: ER-positive (ER+; regardless of PR/HER2 status), ER-negative (ER-; regardless of PR/HER2 status), HER2-positive (HER2+; negative ER/PR), and triple-negative (TN; negative ER/PR/ HER2)[19].

Hypothesis tests were 2-sided (unless otherwise specified) with a significance threshold of 0.05. Statistical analyses were performed using Comprehensive Meta-Analysis Version 3 (Biostat, Englewood, NJ 2013) and R Version 4.0.2.

## 3 RESULTS

### 3.1 Literature search

Initial searches of the databases yielded 10,773 publications. Assessment of references in relevant reviews[7,10–14] yielded an additional 404 publications. After removing duplicates and article types other than research articles, 3,893 publications remained. After removing papers on the basis of title (n=3,150) or abstract (n=488) and excluding reports (n = 23) which were inaccessible online, were conference abstracts with no full text, or had titles or abstracts that do not correspond to those indexed by the search databases, the full text of 232 reports that could be retrieved were reviewed. 70 studies that met eligibility criteria were included in this review and meta-analysis. 2 additional cross-sectional studies assessing molecular subtypes at the time of BCa diagnosis were included in the review but not the meta-analysis. Figure 1 shows the PRISMA flowchart for the study selection process.

**FIGURE 1.**
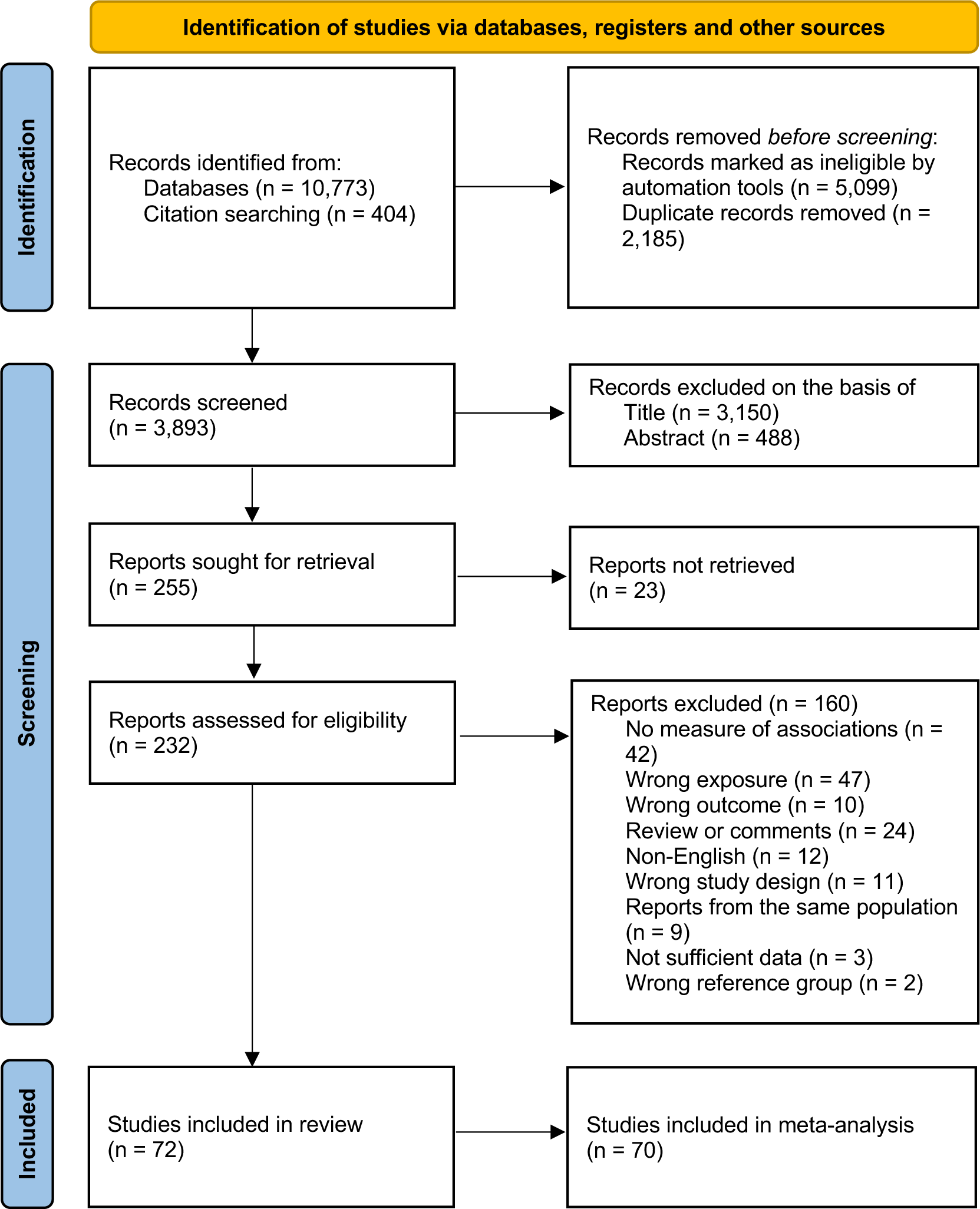
Flow diagram of study selection.

### 3.2 Study description and quality assessment

Descriptive data for the selected studies are presented in Table 1. Of the 72 included studies, 46 were cohort studies, 24 were case-control studies, and the other 2 were cross-sectional studies evaluating BCa molecular subtypes. 24 studies were conducted in Asia, 24 were conducted in the Americas, 21 were conducted in Europe, and 3 were conducted in Australia. Population sizes varied substantially, ranging from 110 to 10,827,079 participants. The slim majority of studies assessed DM based on medical reports (n=39), with some studies adopting a combination of self-report and clinical identification (n=4). 10 studies differentiated T1D from T2D. With the exception of 5 studies, BCa was identified from clinical or cancer registry records. All studies adjusted for age, 36 adjusted for BMI, and 24 adjusted for menopausal status. 52 out of 70 rated studies were deemed medium to high quality, including 34 cohort and 18 case-control studies. The 2 studies that specifically investigated BCa molecular subtypes were not rated due to their cross-sectional design.

**TABLE 1.**
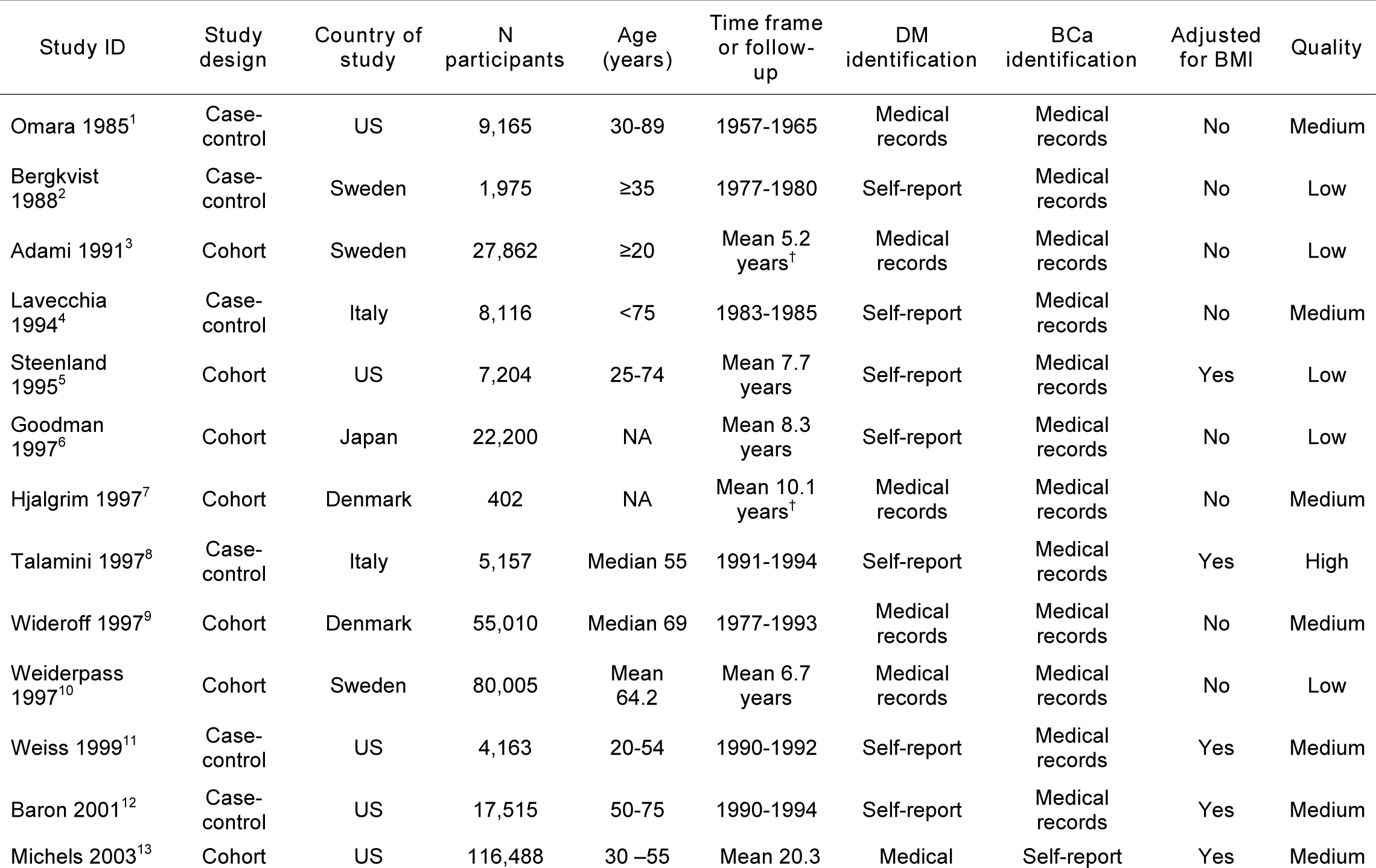
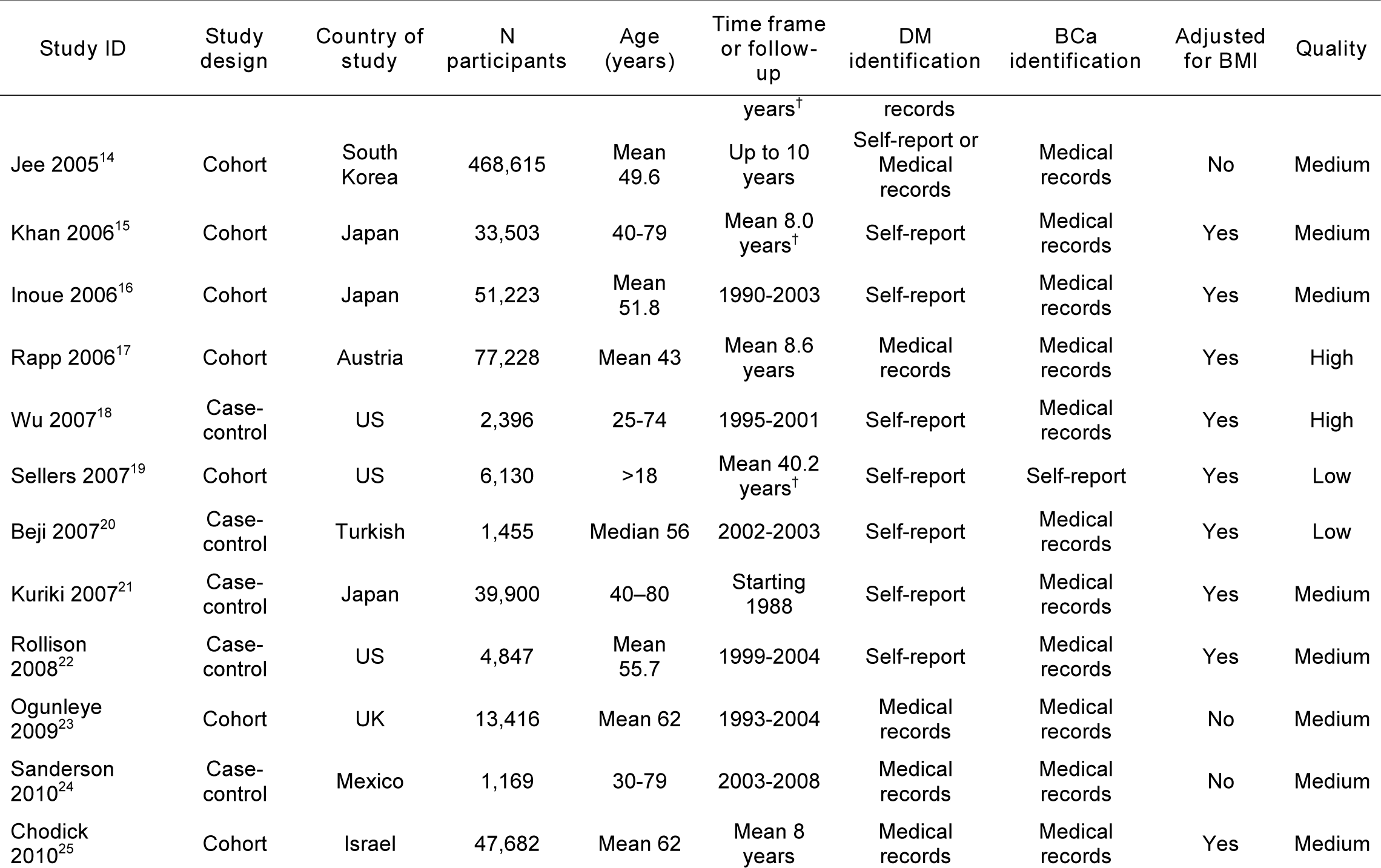
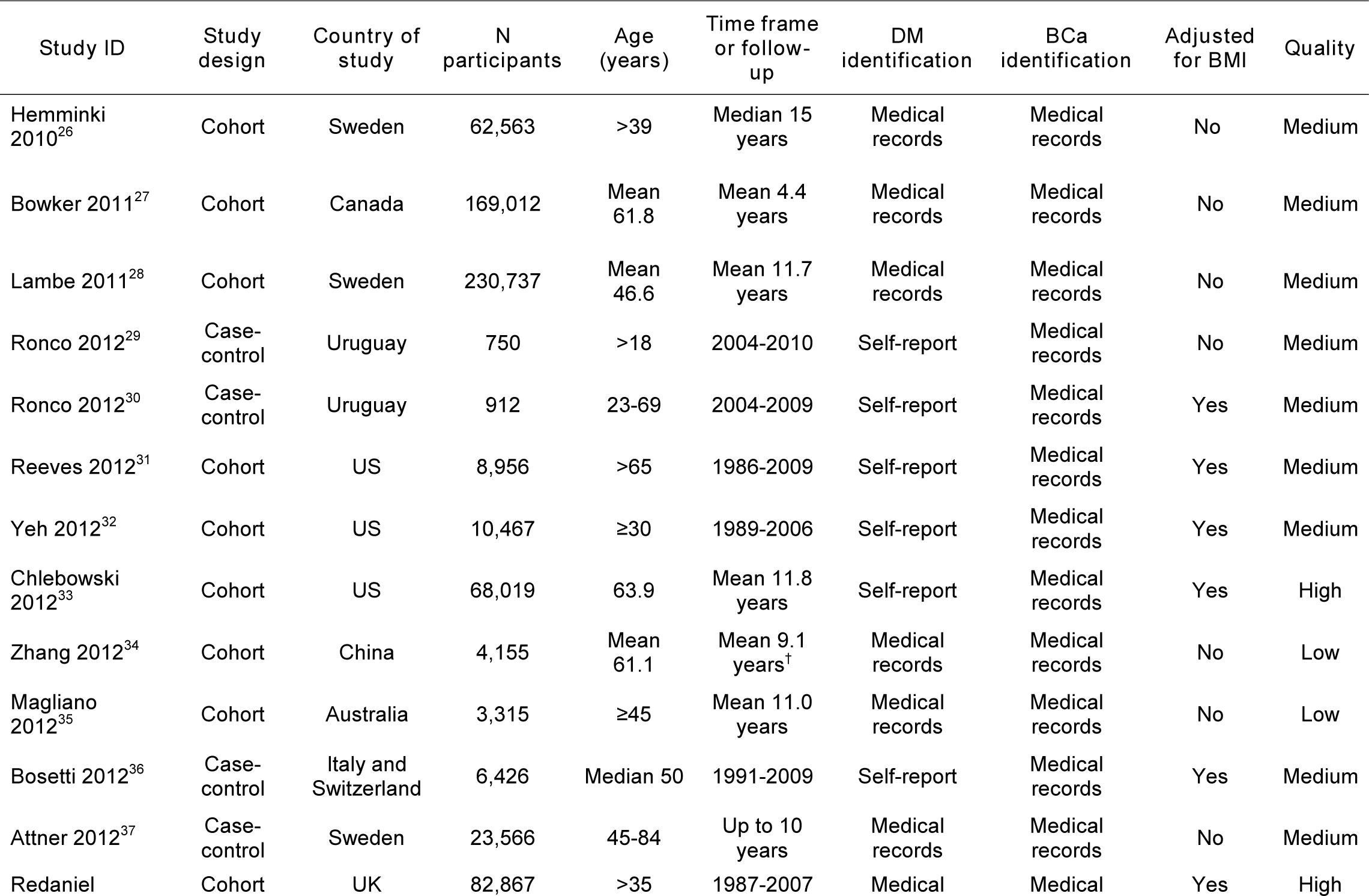
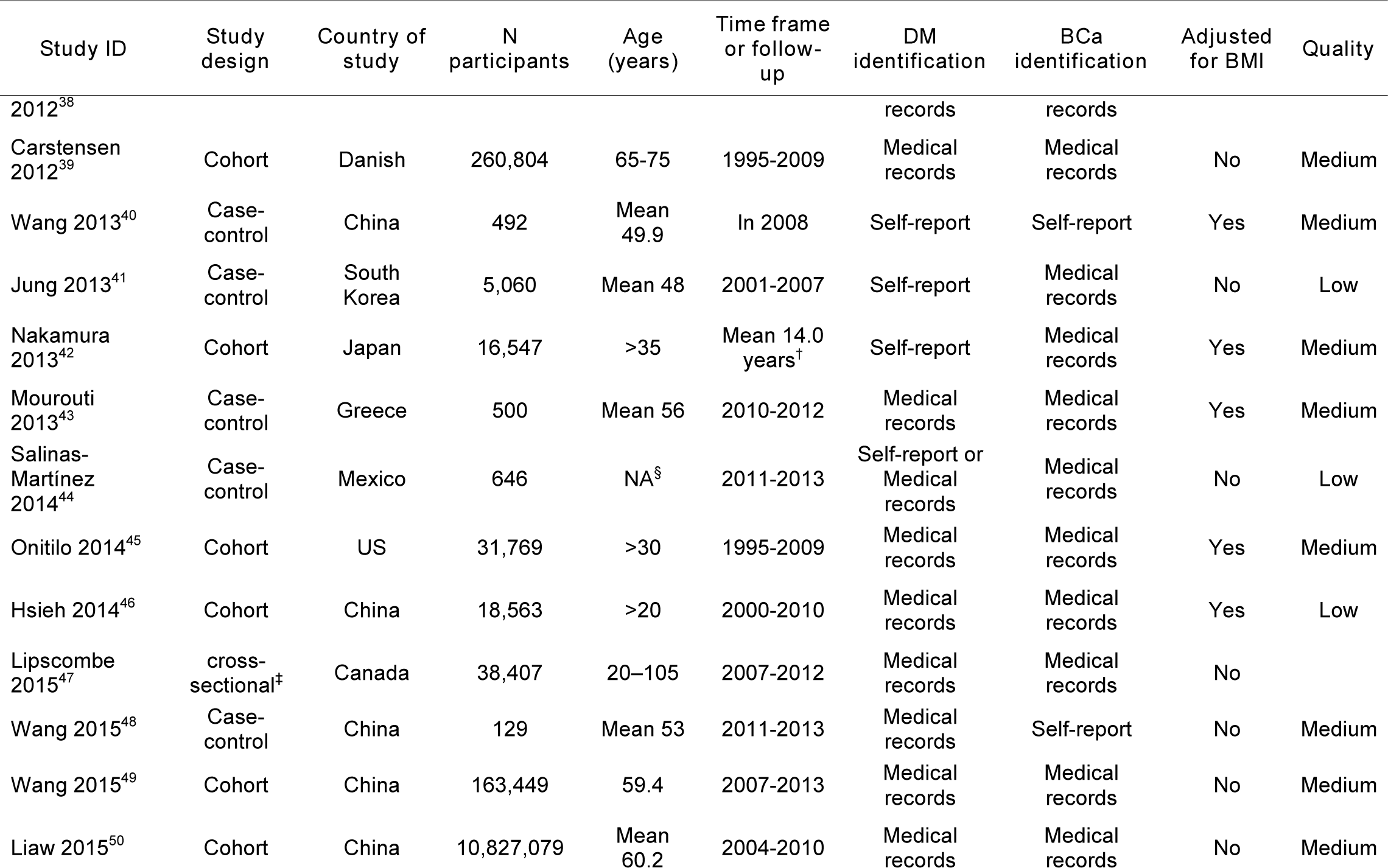
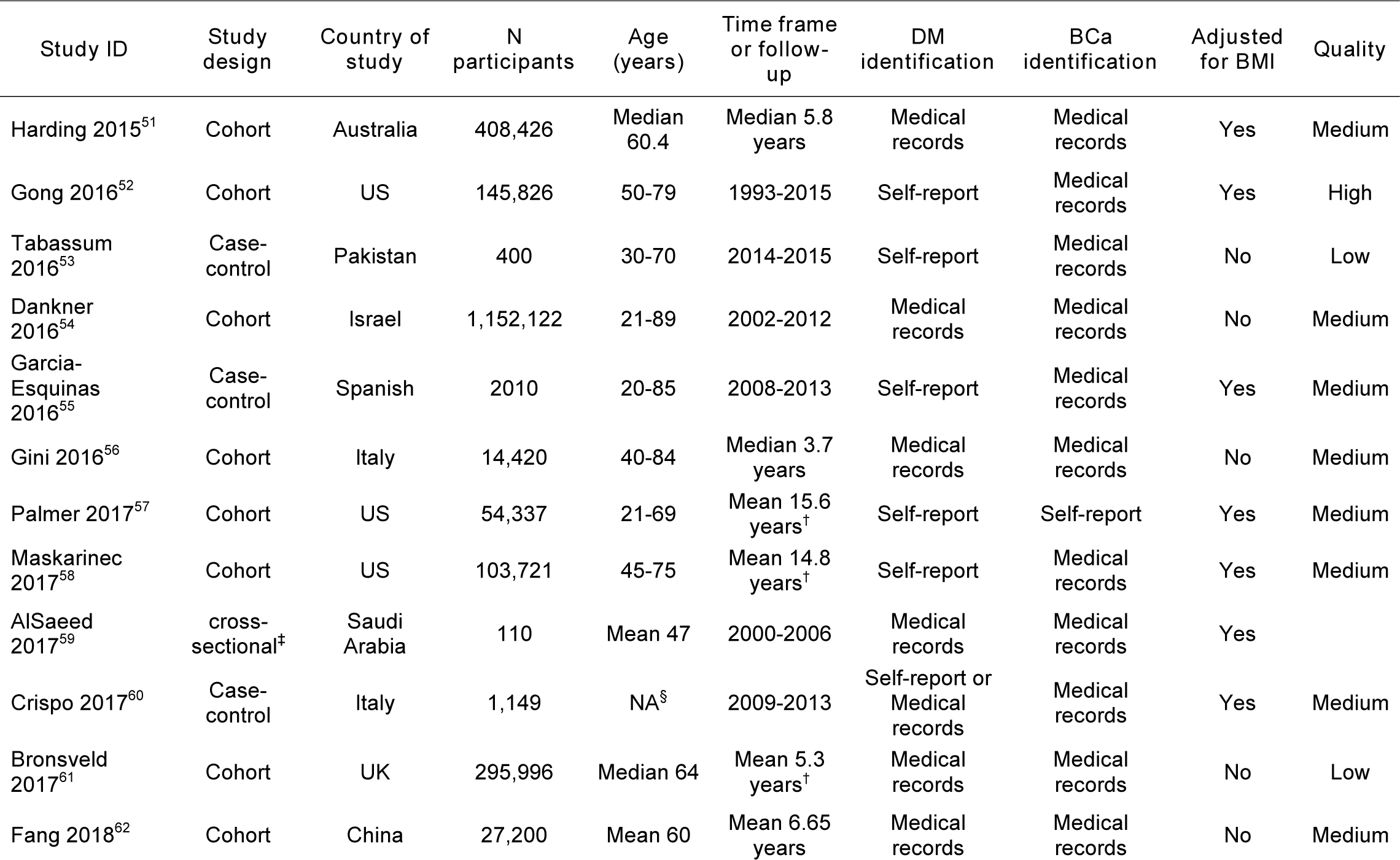
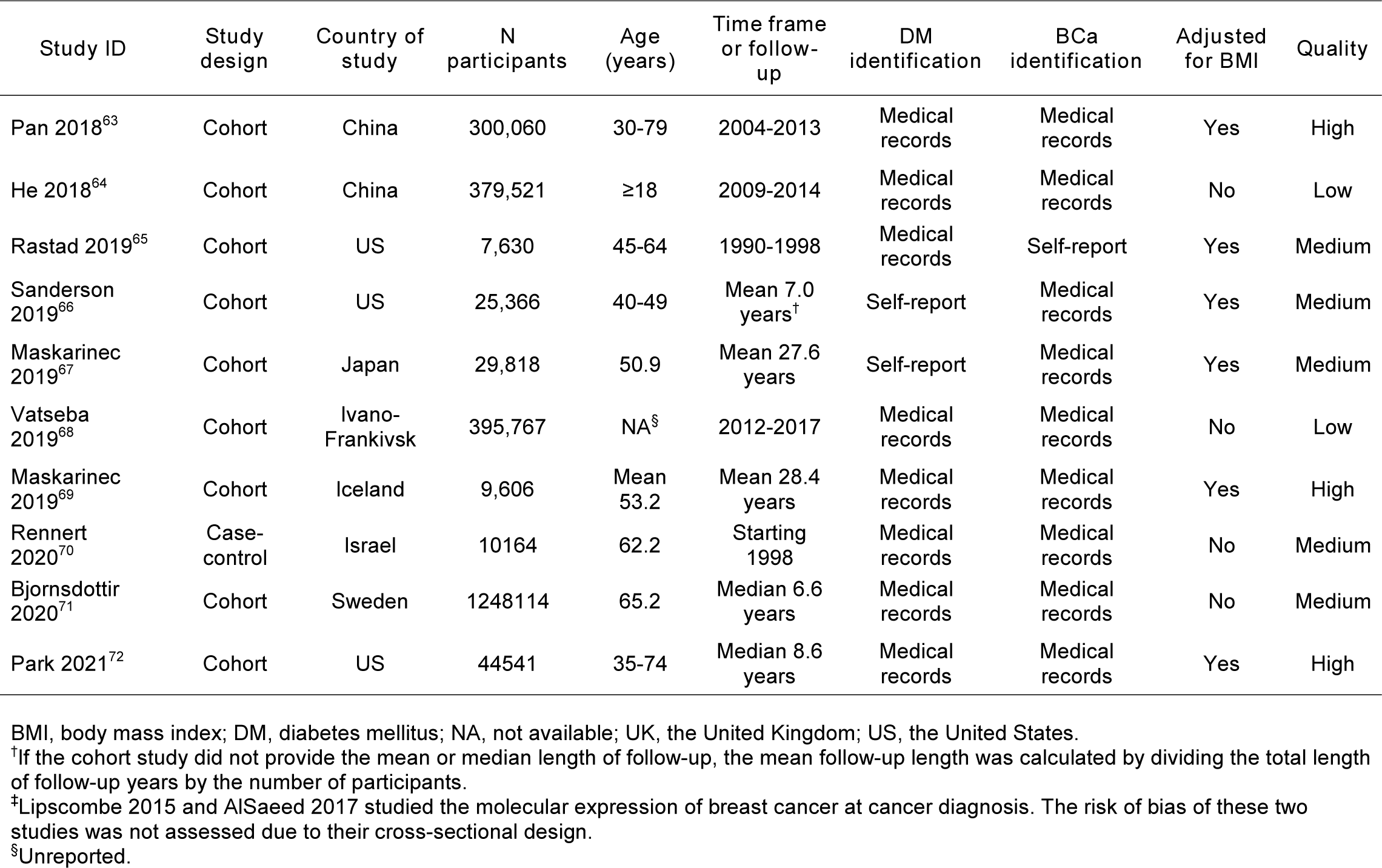
Characteristics of studies included in the systematic review

### 3.3 Meta-analysis results

2 studies were not included in the meta-analysis because they were cross-sectional and only assessed molecular subtypes at the time of BCa diagnosis. The meta-analysis of 70 studies evaluating the association between DM and BCa risk overall demonstrated a 20% increased risk of BCa among women with DM (RR=1.20, 95% CI: 1.11-1.29; Figure 2). Heterogeneity among the studies was substantial (I^2^=97.9%, P- heterogeneity<0.001). Leave-one-out sensitivity analyses revealed that the findings from Dankner 2016[20] were especially influential; excluding this study produced the most attenuated summary estimate (RR=1.14, 95% CI: 1.09-1.19, I^2^=88.7%, P- heterogeneity<0.001; Supplementary Figure S1). The temporal cumulative meta-analysis demonstrated that the summary estimate reached statistical significance in 2001 and remained fairly constant and significant after 2005 (Supplementary Figure S2). Meta-analysis of the 9 high-quality (RR=1.12, 95% CI: 1.02-1.22, I^2^=53.0%, P- heterogeneity=0.03; Table 2) and, separately, 45 medium-quality (RR=1.15, 95% CI: 1.05-1.27, I^2^=98.4%, P-heterogeneity<0.001) studies yielded slightly attenuated summary estimates. The summary RR for the 16 low-quality studies was 1.42 (95% CI: 1.08-1.86, I^2^=96.6%, P-heterogeneity<0.001).

**FIGURE 2.**
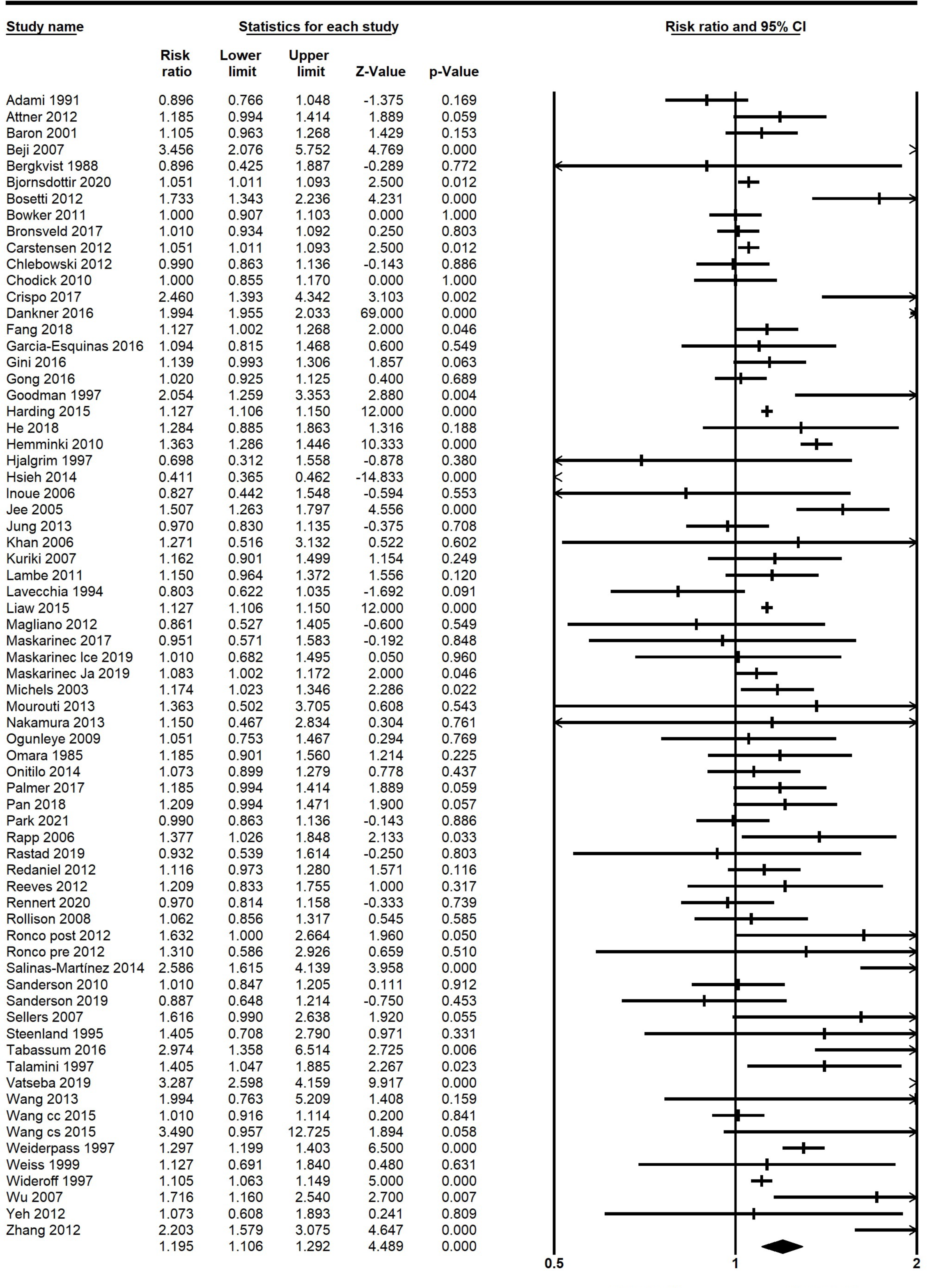
Forest plot of the association between diabetes and breast cancer risk.

**TABLE 2.** Summary estimates of the associations between diabetes and breast cancer risk across subgroups defined by study quality, study design, study region, diabetes ascertainment, adjustment for BMI, year of publication, and menopausal status.

| Subgroup | N studies | Summary estimate | 95% CI | I<sup>2</sup> | P |
| --- | --- | --- | --- | --- | --- |
| All studies | 70 | 1.20 | 1.11-1.29 | 97.9% | <0.001 |
| Study quality |  |  |  |  |  |
| High | 9 | 1.12 | 1.02-1.22 | 53.0% | 0.03 |
| Medium | 45 | 1.15 | 1.05-1.27 | 98.4% | <0.001 |
| Low | 16 | 1.42 | 1.08-1.86 | 96.6% | <0.001 |
| Study design |  |  |  |  |  |
| Cohort | 46 | 1.15 | 1.05-1.27 | 98.6% | <0.001 |
| Case-control | 24 | 1.26 | 1.13-1.40 | 73.1% | <0.001 |
| Study region |  |  |  |  |  |
| Asia | 23 | 1.34 | 1.11-1.63 | 99.1% | <0.001 |
| Americas | 23 | 1.10 | 1.03-1.17 | 40.8% | 0.02 |
| Europe | 21 | 1.13 | 1.06-1.21 | 83.4% | <0.001 |
| Australia | 3 | 1.15 | 0.99-1.33 | 32.0% | 0.23 |
| Year of publication |  |  |  |  |  |
| Before 2012 | 28 | 1.18 | 1.09-1.26 | 77.6% | <0.001 |
| In/after 2012 | 42 | 1.20 | 1.08-1.33 | 98.7% | <0.001 |
| DM ascertainment |  |  |  |  |  |
| Self-report | 30 | 1.18 | 1.09-1.29 | 62.0% | <0.001 |
| Medical records | 37 | 1.14 | 1.03-1.26 | 98.9% | <0.001 |
| Both | 3 | 2.00 | 1.33-3.01 | 68.7% | 0.04 |
| Adjustment for BMI |  |  |  |  |  |
| No | 31 | 1.21 | 1.08-1.35 | 98.9% | <0.001 |
| Yes | 39 | 1.19 | 1.06-1.32 | 89.7% | <0.001 |
| Adjustment for parity |  |  |  |  |  |
| No | 56 | 1.15 | 1.06-1.26 | 98.3% | <0.001 |
| Yes | 14 | 1.39 | 1.18-1.62 | 79.7% | <0.001 |
| Menopausal status<sup>†</sup> |  |  |  |  |  |
| Pre | 15 | 0.95 | 0.85-1.05 | 0.00% | 0.78 |
| Post | 31 | 1.12 | 1.07-1.17 | 65.5% | <0.001 |

Abbreviations: BMI, body mass index; CI, confidence interval; DM, diabetes mellitus.
<sup>†</sup>The numbers of studies in the subgroups defined by menopausal status do not sum up to 70 because only a subset of the studies assessed pre- and post-menopausal breast cancer separately.

Slight publication bias was detected by the funnel plot (Supplementary Figure S3A) and Begg’s test (1-tailed P<0.001), but not by Egger’s test (1-tailed P=0.11). When using trim and fill analysis to measure publication bias, no studies were hypothetically missing, and the summary estimate was therefore unchanged.

The results for subgroup analyses are presented in Table 2. The association estimate summarizing the 46 cohort studies remained statistically significant (RR=1.15, 95% CI: 1.05-1.27), despite 28 studies having reported null findings. The heterogeneity among cohort studies was still considerable (I^2^=98.6%, P-heterogeneity <0.001), but no publication bias was found by the funnel plot or Egger’s test (1-tailed P=0.11). Begg’s test did indicate heterogeneity (1-tailed P <0.001) (Supplementary Figure S3B). Meta-analysis of the 7 cohort studies of high quality showed more homogeneous results (I^2^=23.1%, P-heterogeneity=0.25), with only one study demonstrating a significant association. The summary estimate was not statistically significant (RR=1.06, 95% CI: 0.99-1.14).

The 24 case-control studies showed a significantly increased BCa risk among women with DM (RR=1.26, 95% CI: 1.13-1.40), though 14 studies found no association. Case-control studies demonstrated less heterogeneity than cohort studies (I^2^=73.1%, P-heterogeneity<0.001). However, the funnel plot demonstrated significant asymmetry, wherein nearly all small studies with less than average estimates were missing (Supplementary Figure S3C). After imputing 9 hypothetically missing association estimates to the bottom left of the funnel plot, the summary estimate decreased remarkably and was no longer statistically significant (RR=1.05, 95% CI: 0.93-1.19). The 2 case-control studies that were deemed high quality reported ORs of 1.40 (95% CI: 1.00-1.96) and 1.70 (95% CI: 1.15-2.54), respectively. The pooled estimate from 19 medium- to high-quality case-control studies was 1.17 (95% CI: 1.07-1.29, I^2^=59.8%, P- heterogeneity<0.001).

Among regions, the largest association estimate between DM and BCa risk was found in Asia (RR=1.34, 95% CI: 1.11-1.63, I^2^=99.1%, P-heterogeneity<0.001, n=23), followed by Europe (RR=1.13, 95% CI: 1.06-1.21, I^2^=83.4%, P-heterogeneity<0.001, n=21), and the Americas (RR=1.10, 95% CI: 1.03-1.17, I^2^=40.8%, P-heterogeneity=0.02, n=23). The 3 Australian studies showed homogeneous findings, and though the summary association estimate was comparable to that for European studies, the combined results were not statistically significant (RR=1.15, 95% CI: 0.99-1.33, I^2^=32.0%, P-heterogeneity=0.23). Estimates for studies published before 2012 (RR=1.18, 95% CI: 1.09-1.26, I^2^=77.6%, P-heterogeneity<0.001, n=28) versus in/after 2012 (RR=1.20, 95% CI: 1.08-1.33, I^2^=98.7%, P-heterogeneity<0.001, n=42) were essentially comparable. Methods of DM identification did not materially influence association estimates (self-report RR=1.18, 95% CI: 1.09-1.29, I^2^=62.0%, P- heterogeneity<0.001, n=30 versus medical records RR=1.14, 95% CI: 1.03-1.26, I^2^=98.9%, P-heterogeneity<0.001, n=37). Studies that did not adjust for BMI yielded an equivalent summary estimate (RR=1.21, 95% CI: 1.08-1.35, I^2^=98.9%, P- heterogeneity<0.001, n=31) to studies controlling for BMI (RR=1.19, 95% CI: 1.06-1.32, I^2^=89.7%, P-heterogeneity<0.001, n=39). 56 studies not accounting for parity yielded a summary estimate of 1.15 (95% CI: 1.06-1.26, I^2^=98.3%, P-heterogeneity<0.001), while the summary estimate of the remaining 14 studies was 1.39 (95% CI: 1.18-1.62, I^2^=79.7%, P-heterogeneity<0.001). The meta-regressions did not suggest heterogeneity across any subgroups.

31 studies, including 11 cohort studies, measured BCa risk in postmenopausal women, for whom the summary RR was 1.12 (95% CI: 1.07-1.17, I^2^=65.5%, P-heterogeneity<0.001). 15 studies reported estimates in premenopausal women, and none of them found significant associations. We also did not observe an overall association for premenopausal BCa, and the between-study heterogeneity was negligible (RR=0.95, 95% CI: 0.85-1.05, I^2^=0.00%, P-heterogeneity=0.78).

13 studies (including the two cross-sectional studies that were not included in the other meta-analyses) evaluated associations with BCa molecular subtypes. All of them presented results for ER+ BCa, and the summary RR was 1.09 (95% CI: 1.00-1.20, I^2^=52.4%, P-heterogeneity=0.01; Figure 3A). Analysis based on 11 studies showed an increased risk of ER- BCa associated with DM (RR=1.16, 95% CI: 1.04-1.30, I^2^=0.00%, P-heterogeneity=0.57; Figure 3B). The summary estimate for 3 studies that reported data for HER2+ BCa did not reveal a statistically significant association, and the confidence interval was especially wide (RR=1.21, 95% CI: 0.52-2.82, I^2^=67.5%, P-heterogeneity=0.05; Figure 3C). The 5 studies that assessed the risk of TN BCa showed a significantly elevated risk related to DM (RR=1.41, 95% CI: 1.01-1.96, I^2^=49.7%, P-heterogeneity=0.09; Figure 3D).

**FIGURE 3.**
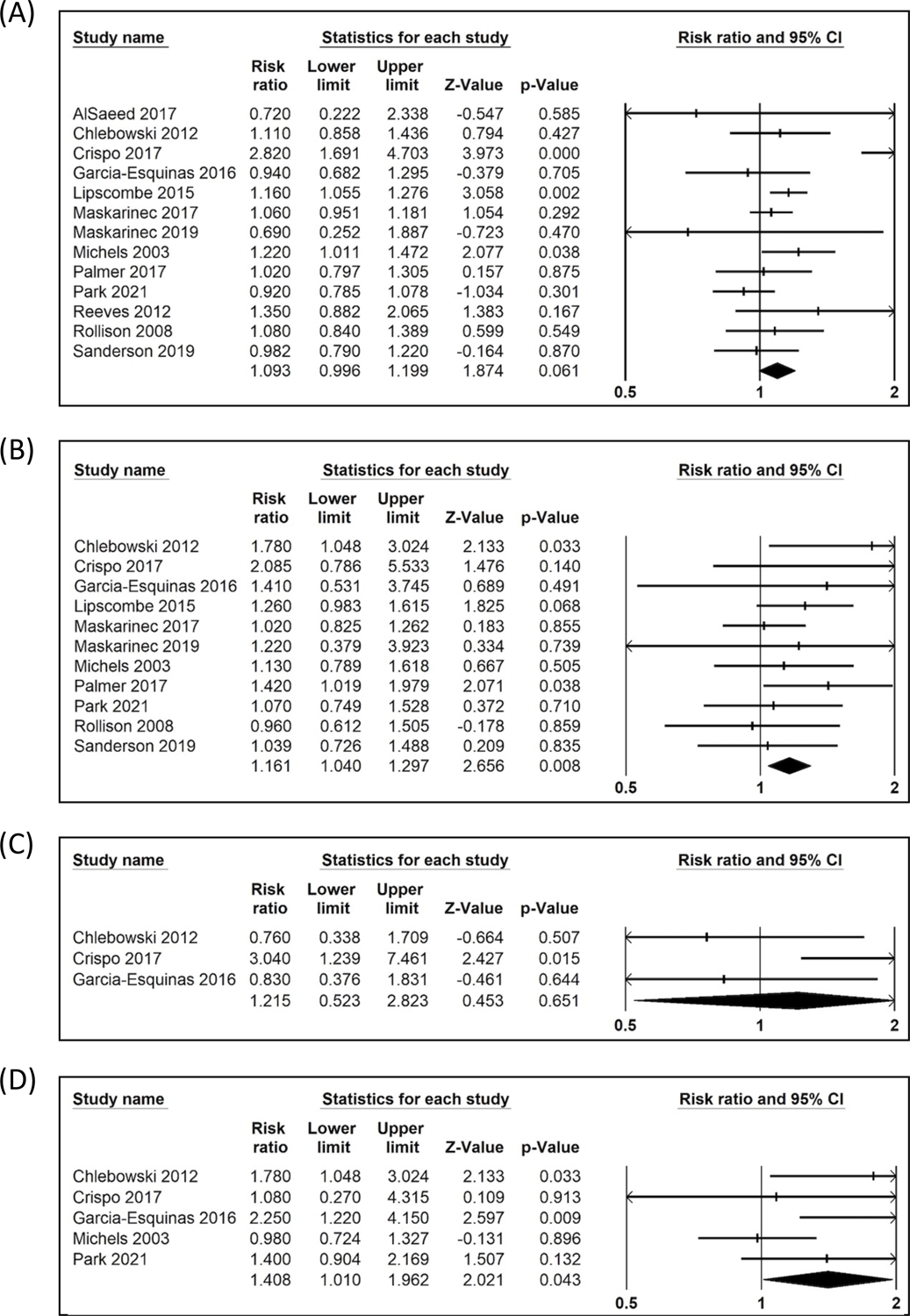
Forest plots for the associations of diabetes with (A) estrogen receptor-positive breast cancer, (B) estrogen receptor-negative breast cancer, (C) triple-negative breast cancer, and (D) human epidermal growth factor 2-positive breast cancer.

## 4 DISCUSSION

In this review of evidence regarding a possible association between DM and BCa risk, we observed a positive relationship in many studies, even while more than half of studies reported null results. In the overall meta-analysis including 70 cohort and case-control studies, we identified a 20% higher risk of BCa in women with DM. The association was more modest but still statistically significant when restricting to high-quality studies.

Our findings of increased BCa risk in women with DM align with conclusions from previous reviews[7,10–14]. Wolf et al. (2005) was the first to summarize the association[11]. Their findings based on 6 cohort and 4 case-control studies suggested that T2D, but not T1D, may be associated with 10-20% excess risk of BCa. The reviews and meta-analyses that have been published since have reported similar summary estimates[7,10,12–14]. The largest review, which included 40 observational studies, was conducted by Hardefeldt et al. in 2012. It demonstrated a 20% increased risk of BCa associated with DM[7]. In contrast to our primary analyses, Hardefeldt et al. included cross-sectional studies and gestational DM. We largely focused on cohort and case-control studies in an effort to consider temporality. We did not evaluate gestational DM because its pathogenesis differs from that of other DM, which could render its impact on breast tumorigenesis distinct. A recent meta-analysis assessed the relationship of gestational DM with BCa risk with null results[21,22].

Several biological mechanisms for BCa initiation in diabetic patients have been postulated. DM is characterized by hyperglycemia and hyperinsulinemia. Regarding the former, excessive glucose could favor the selection of malignant cell clones[23] and/or induce DNA damage, downregulate the expression of antioxidants[24], and increase the generation of reactive oxygen species[25,26]. Regarding the latter, increased insulin levels resulting from resistance to endogenous insulin or insulin treatment could directly promote BCa cell proliferation and inhibit cell apoptosis or indirectly mediate growth-promoting actions by increasing hepatic insulin-like growth factor-1 (IGF-1) production^17–19^. The mechanical pathways of fat-induced inflammatory cytokines and altered levels of sex hormone and sex hormone-binding globulin may also play a role[27,28]. Preclinical data suggest that use of the first-line antidiabetic therapy metformin could be beneficial for BCa prevention because it directly suppresses cell proliferation through the activation of AMPK and regulation of the mTOR signaling pathway or indirectly improves hyperglycemia, hyperinsulinemia, and inflammation[29–31]. However, epidemiologic evidence regarding the relationship between metformin and BCa prevention has been inconsistent[32].

Despite the overall positive association between DM and BCa that we identified, heterogeneity across studies was substantial. Population composition, study design, study region, and covariate adjustment all impacted the findings. In particular, our meta-analysis of cohort studies yielded a more modest summary estimate than our meta-analysis of case-control studies. Publication bias, however, may not have been negligible among case-control studies, and imputation of potentially missing estimates from individual studies yielded a summary estimate that was attenuated and non-significant.

We observed a 34% increased risk of BCa in women with DM in studies conducted in Asia, whereas studies conducted in Western populations demonstrated smaller associations. We note that heterogeneity was highest in the meta-analysis of studies conducted in Asia. To explore heterogeneity among such studies, we ran *a posteriori* meta-regression analyses that demonstrated a significant difference between studies that did versus did not adjust for BMI. 13 studies that did not adjust for BMI indicated a significant 53% increased risk of BCa among women with DM, while 10 studies that did adjust for BMI did not yield a significant summary association.

Though adjustment for BMI was consequential for studies in Asia, BMI-adjusted studies overall had a comparable summary estimate to studies that did not adjust for BMI. These results suggest that DM may be a risk factor for BCa independent of obesity. Nonetheless, obesity and DM might trigger similar pathogenic mechanisms of BCa development, including metabolic changes, inflammation, insulin resistance and increased IGF-1, and altered antitumor immunity[33]. Parity, and particularly multiple pregnancies, has been linked with an increased risk of T2DM but is associated with a lower risk of BCa[34,35]. Ignoring parity when evaluating the relationship between T2M and BCa could result in underestimations of the true association. Indeed, we found that studies that adjusted for parity yielded slightly stronger association estimates.

BCa is a heterogeneous disease with varied molecular features, but the association between DM and BCa molecular subtypes is uncertain. Biological evidence suggests that hyperinsulinemia could induce expression and increase the binding ability of ER[36]. By contrast, metformin may repress the expression of ER and ER target genes[37]. DM-induced changes in levels of bioavailable sex hormones, such as increasing levels of estrogen and androgen and decreasing levels of progesterone[36], could also impact receptor expression. One meta-analysis found non-significant associations between DM and ER, PR, and HER2 status, respectively, by comparing hormone receptor negativity to their positive counterparts[38]. In our review, we found that DM was most strongly associated with TN BCa, which is among the most aggressive types of disease.

We were not able to combine findings regarding associations between DM duration and BCa risk since studies had different DM duration categories. However, multiple, though not all, studies that assessed the temporal trend of the association between DM and BCa indicated that the relationship was strongest during the time period closely following DM diagnosis[39–45]. It is possible that women with newly diagnosed DM have more frequent contact with health care providers and are thus more likely to uncover BCa. Neglecting this detection bias may lead to overestimation of BCa risk at the time of DM diagnosis[46]. However, screening bias is unlikely to account for the excess risk of BCa observed up to 10 years following DM diagnosis[42,45,47,48], suggesting that DM may play a real etiologic role in BCa development.

Our review expands upon findings from previous reviews in several important ways. First, we assessed the risk of bias for each study. As such, we were able to combine findings from only high-quality studies, which were least likely to be affected by selection bias, confounding, and reverse causation. Second, we identified and adjusted for potential missingness of case-control studies with small sample sizes and non-significant results. Third, our study summarized existing evidence addressing possible associations between DM and molecular subtypes of BCa. Nevertheless, there are some limitations of this review worth noting. Many of the contributing studies were susceptible to selection bias, most often related to restriction to older ages, so as to maximize the number of DM and/or BCa cases. In addition, most studies were unable to differentiate between T1D and T2D. Because T1D and T2D may differ in etiology, patient demographics, and treatment regimens, the pathogenesis of BCa could be different in these two patient groups. It is reasonable to assume that the results of this review largely concern T2D given that more than 95% of DM cases are estimated to be T2D[49], and in many studies, the median/mean age at DM diagnosis was above 40 years. Finally, most included studies did not report information on DM therapy, which could mediate any relationship between DM and BCa risk.

In conclusion, this updated systematic review and meta-analysis confirms prior findings of an increased risk of BCa in women with DM. In addition, our results suggest that women with DM may have a higher risk of diagnosis with an aggressive molecular subtype of BCa compared to women without DM. Further high-quality evidence is needed to verify the relationship between DM and BCa receptor expression, understand the underlying mechanisms of breast carcinogenesis in women with DM, and eventually provide evidence-based recommendations for BCa prevention and treatment.

## Supporting information

Search appendix

## Data Availability

All data produced in the present study are available upon reasonable request to the authors

## ACKNOWLEDGEMENTS

We gratefully acknowledge Dr. Chiung-Yu Huang and Dr. Thomas Newman for their critical feedback.

## SUPPORTING INFORMATION

SUPPLEMENTARY FIGURE S1. Leave-one-out sensitivity meta-analyses of 70 studies assessing the association between diabetes and breast cancer risk.

SUPPLEMENTARY FIGURE S2. Cumulative temporal meta-analysis of 70 studies assessing the association between diabetes and breast cancer risk.

SUPPLEMENTARY FIGURE S3. Funnel plots of observed and imputed studies using trim-and-fill method for assessing publication bias among (A) all 70 studies, (B) 46 cohort studies, and (C) 24 case-control studies.TABLE 2. Summary estimates of the associations between diabetes and breast cancer risk across subgroups defined by study

## Notes

**FUNDING** JMC receives funding from the Cancer League Foundation. REG is supported by a Young Investigator Award from the Prostate Cancer Foundation.

### Competing Interest Statement

JMC declares that her husband is a full-time employee of Adela, Inc. and a prior employee of GRAIL (within the past three years).

### Funding Statement

JMC receives funding from the Cancer League Foundation. REG is supported by a Young Investigator Award from the Prostate Cancer Foundation.

