## Supplementary Figure 1 for "Diabetes and Incidence of Breast Cancer and its Molecular Subtypes: A Systematic Review and Meta-Analysis"

| Study name | Statistics with study removed | | | | |
|---|---|---|---|---|---|
| | Point | Lower limit | Upper limit | Z-Value | p-Value |
| Adami 1991 | 1.202 | 1.111 | 1.300 | 4.580 | 0.000 |
| Attner 2012 | 1.196 | 1.105 | 1.294 | 4.448 | 0.000 |
| Baron 2001 | 1.197 | 1.106 | 1.295 | 4.477 | 0.000 |
| Beji 2007 | 1.182 | 1.093 | 1.278 | 4.188 | 0.000 |
| Bergkvist 1988 | 1.198 | 1.108 | 1.295 | 4.524 | 0.000 |
| Bjornsdottir 2020 | 1.199 | 1.107 | 1.300 | 4.445 | 0.000 |
| Bosetti 2012 | 1.188 | 1.098 | 1.285 | 4.303 | 0.000 |
| Bowker 2011 | 1.200 | 1.109 | 1.298 | 4.524 | 0.000 |
| Bronsveld 2017 | 1.199 | 1.108 | 1.298 | 4.514 | 0.000 |
| Carstensen 2012 | 1.199 | 1.107 | 1.300 | 4.445 | 0.000 |
| Chlebowski 2012 | 1.200 | 1.109 | 1.298 | 4.531 | 0.000 |
| Chodick 2010 | 1.199 | 1.109 | 1.297 | 4.526 | 0.000 |
| Crispo 2017 | 1.187 | 1.098 | 1.284 | 4.296 | 0.000 |
| Dankner 2016 | 1.141 | 1.091 | 1.192 | 5.851 | 0.000 |
| Fang 2018 | 1.197 | 1.106 | 1.295 | 4.465 | 0.000 |
| Garcia-Esquinas 2016 | 1.197 | 1.107 | 1.295 | 4.487 | 0.000 |
| Gini 2016 | 1.197 | 1.106 | 1.295 | 4.463 | 0.000 |
| Gong 2016 | 1.199 | 1.108 | 1.297 | 4.513 | 0.000 |
| Goodman 1997 | 1.188 | 1.099 | 1.285 | 4.316 | 0.000 |
| Harding 2015 | 1.202 | 1.103 | 1.310 | 4.182 | 0.000 |
| He 2018 | 1.194 | 1.104 | 1.292 | 4.433 | 0.000 |
| Hemminki 2010 | 1.193 | 1.102 | 1.292 | 4.343 | 0.000 |
| Hjalgrim 1997 | 1.199 | 1.109 | 1.297 | 4.561 | 0.000 |
| Hsieh 2014 | 1.217 | 1.129 | 1.312 | 5.119 | 0.000 |
| Inoue 2006 | 1.199 | 1.109 | 1.297 | 4.549 | 0.000 |
| Jee 2005 | 1.191 | 1.100 | 1.288 | 4.344 | 0.000 |
| Jung 2013 | 1.200 | 1.109 | 1.298 | 4.541 | 0.000 |
| Khan 2006 | 1.195 | 1.105 | 1.292 | 4.468 | 0.000 |
| Kuriki 2007 | 1.196 | 1.106 | 1.294 | 4.462 | 0.000 |
| Lambe 2011 | 1.196 | 1.106 | 1.294 | 4.462 | 0.000 |
| Lavecchia 1994 | 1.203 | 1.112 | 1.301 | 4.616 | 0.000 |
| Liaw 2015 | 1.202 | 1.103 | 1.310 | 4.182 | 0.000 |
| Magliano 2012 | 1.200 | 1.109 | 1.297 | 4.555 | 0.000 |
| Maskarinec 2017 | 1.198 | 1.108 | 1.296 | 4.526 | 0.000 |
| Maskarinec Ice 2019 | 1.198 | 1.108 | 1.296 | 4.514 | 0.000 |
| Maskarinec Ja 2019 | 1.198 | 1.107 | 1.297 | 4.474 | 0.000 |
| Michels 2003 | 1.196 | 1.105 | 1.294 | 4.449 | 0.000 |
| Mourouti 2013 | 1.195 | 1.105 | 1.292 | 4.463 | 0.000 |
| Nakamura 2013 | 1.196 | 1.106 | 1.293 | 4.482 | 0.000 |
| Ogunleye 2009 | 1.198 | 1.107 | 1.295 | 4.502 | 0.000 |
| Omara 1985 | 1.196 | 1.105 | 1.293 | 4.455 | 0.000 |
| Onitilo 2014 | 1.198 | 1.107 | 1.296 | 4.494 | 0.000 |
| Palmer 2017 | 1.196 | 1.105 | 1.294 | 4.448 | 0.000 |
| Pan 2018 | 1.195 | 1.105 | 1.293 | 4.441 | 0.000 |
| Park 2021 | 1.200 | 1.109 | 1.298 | 4.531 | 0.000 |
| Rapp 2006 | 1.193 | 1.103 | 1.290 | 4.399 | 0.000 |
| Rastad 2019 | 1.198 | 1.108 | 1.296 | 4.528 | 0.000 |
| Redaniel 2012 | 1.197 | 1.106 | 1.295 | 4.472 | 0.000 |
| Reeves 2012 | 1.195 | 1.105 | 1.293 | 4.454 | 0.000 |
| Rennert 2020 | 1.200 | 1.109 | 1.298 | 4.540 | 0.000 |
| Rollison 2008 | 1.198 | 1.107 | 1.296 | 4.499 | 0.000 |
| Ronco post 2012 | 1.191 | 1.101 | 1.288 | 4.378 | 0.000 |
| Ronco pre 2012 | 1.195 | 1.105 | 1.292 | 4.460 | 0.000 |
| Salinas-Martínez 2014 | 1.185 | 1.096 | 1.281 | 4.246 | 0.000 |
| Sanderson 2010 | 1.199 | 1.108 | 1.297 | 4.521 | 0.000 |
| Sanderson 2019 | 1.201 | 1.110 | 1.299 | 4.567 | 0.000 |
| Sellers 2007 | 1.191 | 1.102 | 1.289 | 4.381 | 0.000 |
| Steenland 1995 | 1.194 | 1.104 | 1.291 | 4.440 | 0.000 |
| Tabassum 2016 | 1.188 | 1.099 | 1.285 | 4.326 | 0.000 |
| Talamini 1997 | 1.192 | 1.102 | 1.290 | 4.392 | 0.000 |
| Vatseba 2019 | 1.175 | 1.087 | 1.270 | 4.055 | 0.000 |
| Wang 2013 | 1.192 | 1.103 | 1.289 | 4.415 | 0.000 |
| Wang cc 2015 | 1.199 | 1.108 | 1.298 | 4.518 | 0.000 |
| Wang cs 2015 | 1.191 | 1.102 | 1.288 | 4.400 | 0.000 |
| Weiderpass 1997 | 1.194 | 1.103 | 1.292 | 4.383 | 0.000 |
| Weiss 1999 | 1.196 | 1.106 | 1.294 | 4.480 | 0.000 |
| Wideroff 1997 | 1.199 | 1.106 | 1.299 | 4.403 | 0.000 |
| Wu 2007 | 1.190 | 1.100 | 1.287 | 4.342 | 0.000 |
| Yeh 2012 | 1.197 | 1.106 | 1.294 | 4.493 | 0.000 |
| Zhang 2012 | 1.185 | 1.096 | 1.281 | 4.240 | 0.000 |
| | 1.195 | 1.106 | 1.292 | 4.489 | 0.000 |

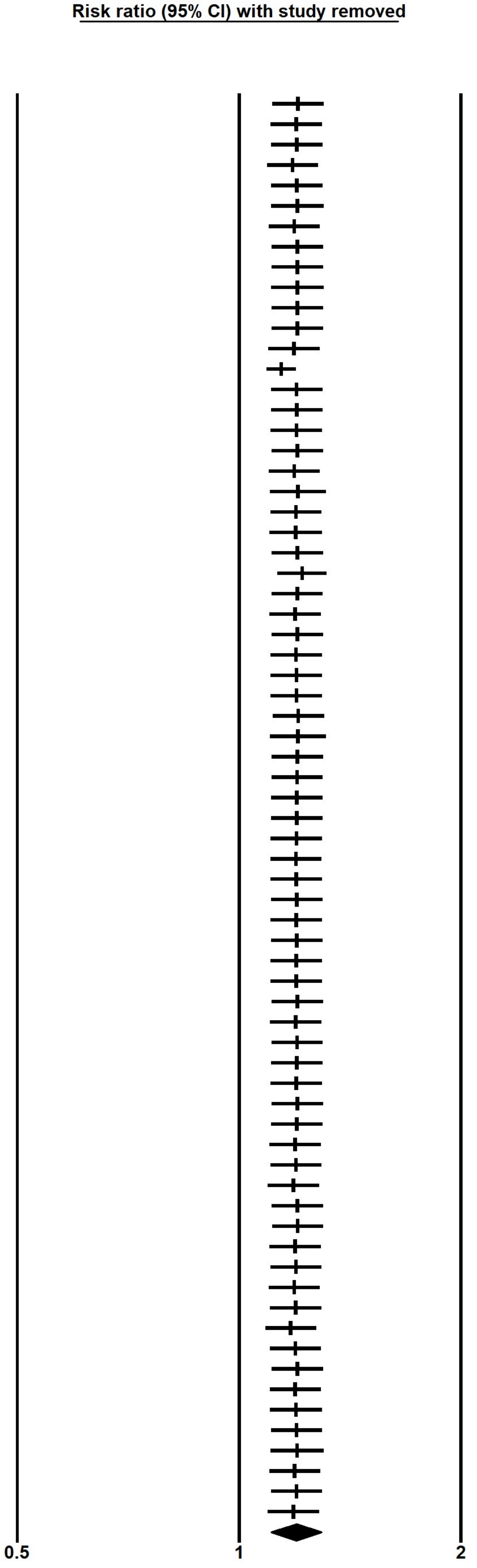
