## Supplementary Figure 2 for "Diabetes and Incidence of Breast Cancer and its Molecular Subtypes: A Systematic Review and Meta-Analysis"

| Study name | Point | Lower limit | Upper limit | Z-Value | p-Value |
|---|---|---|---|---|---|
| Omara 1985 | 1.185 | 0.901 | 1.560 | 1.214 | 0.225 |
| Bergkvist 1988 | 1.146 | 0.886 | 1.483 | 1.039 | 0.299 |
| Adami 1991 | 0.986 | 0.804 | 1.209 | -0.137 | 0.891 |
| Lavecchia 1994 | 0.931 | 0.790 | 1.096 | -0.857 | 0.391 |
| Steenland 1995 | 0.952 | 0.807 | 1.124 | -0.575 | 0.565 |
| Goodman 1997 | 1.080 | 0.846 | 1.380 | 0.618 | 0.536 |
| Hjalgrim 1997 | 1.048 | 0.832 | 1.320 | 0.397 | 0.692 |
| Talamini 1997 | 1.101 | 0.883 | 1.374 | 0.855 | 0.393 |
| Weiderpass 1997 | 1.133 | 0.936 | 1.373 | 1.279 | 0.201 |
| Wideroff 1997 | 1.122 | 0.991 | 1.271 | 1.812 | 0.070 |
| Weiss 1999 | 1.122 | 0.996 | 1.264 | 1.887 | 0.059 |
| Baron 2001 | 1.119 | 1.009 | 1.241 | 2.127 | 0.033 |
| Michels 2003 | 1.125 | 1.027 | 1.233 | 2.526 | 0.012 |
| Jee 2005 | 1.158 | 1.055 | 1.271 | 3.076 | 0.002 |
| Inoue 2006 | 1.151 | 1.049 | 1.262 | 2.976 | 0.003 |
| Khan 2006 | 1.152 | 1.051 | 1.261 | 3.041 | 0.002 |
| Rapp 2006 | 1.163 | 1.065 | 1.270 | 3.353 | 0.001 |
| Beji 2007 | 1.200 | 1.087 | 1.325 | 3.625 | 0.000 |
| Kuriki 2007 | 1.197 | 1.090 | 1.315 | 3.751 | 0.000 |
| Sellers 2007 | 1.207 | 1.100 | 1.324 | 3.965 | 0.000 |
| Wu 2007 | 1.223 | 1.115 | 1.342 | 4.268 | 0.000 |
| Rollison 2008 | 1.211 | 1.109 | 1.323 | 4.263 | 0.000 |
| Ogunleye 2009 | 1.204 | 1.105 | 1.312 | 4.252 | 0.000 |
| Chodick 2010 | 1.188 | 1.095 | 1.289 | 4.141 | 0.000 |
| Hemminki 2010 | 1.202 | 1.110 | 1.302 | 4.529 | 0.000 |
| Sanderson 2010 | 1.190 | 1.102 | 1.286 | 4.430 | 0.000 |
| Bowker 2011 | 1.177 | 1.092 | 1.268 | 4.287 | 0.000 |
| Lambe 2011 | 1.175 | 1.094 | 1.262 | 4.412 | 0.000 |
| Attner 2012 | 1.175 | 1.097 | 1.259 | 4.578 | 0.000 |
| Bosetti 2012 | 1.192 | 1.112 | 1.278 | 4.949 | 0.000 |
| Carstensen 2012 | 1.180 | 1.107 | 1.257 | 5.104 | 0.000 |
| Chlebowski 2012 | 1.170 | 1.100 | 1.245 | 4.979 | 0.000 |
| Magliano 2012 | 1.166 | 1.096 | 1.240 | 4.890 | 0.000 |
| Redaniel 2012 | 1.163 | 1.096 | 1.234 | 4.988 | 0.000 |
| Reeves 2012 | 1.164 | 1.097 | 1.234 | 5.068 | 0.000 |
| Ronco post 2012 | 1.168 | 1.102 | 1.238 | 5.217 | 0.000 |
| Ronco pre 2012 | 1.169 | 1.103 | 1.239 | 5.259 | 0.000 |
| Yeh 2012 | 1.168 | 1.102 | 1.237 | 5.268 | 0.000 |
| Zhang 2012 | 1.184 | 1.116 | 1.256 | 5.602 | 0.000 |
| Jung 2013 | 1.175 | 1.109 | 1.245 | 5.462 | 0.000 |
| Mourouti 2013 | 1.176 | 1.110 | 1.246 | 5.495 | 0.000 |
| Nakamura 2013 | 1.175 | 1.110 | 1.245 | 5.512 | 0.000 |
| Wang 2013 | 1.178 | 1.112 | 1.247 | 5.579 | 0.000 |
| Hsieh 2014 | 1.158 | 1.067 | 1.258 | 3.511 | 0.000 |
| Onitilo 2014 | 1.155 | 1.066 | 1.252 | 3.524 | 0.000 |
| Salinas-Martínez 2014 | 1.171 | 1.080 | 1.269 | 3.840 | 0.000 |
| Harding 2015 | 1.154 | 1.081 | 1.232 | 4.283 | 0.000 |
| Liaw 2015 | 1.139 | 1.078 | 1.203 | 4.681 | 0.000 |
| Wang cc 2015 | 1.134 | 1.075 | 1.196 | 4.620 | 0.000 |
| Wang cs 2015 | 1.136 | 1.077 | 1.198 | 4.688 | 0.000 |
| Dankner 2016 | 1.201 | 1.091 | 1.321 | 3.733 | 0.000 |
| Garcia-Esquinas 2016 | 1.198 | 1.090 | 1.317 | 3.735 | 0.000 |
| Gini 2016 | 1.197 | 1.090 | 1.314 | 3.763 | 0.000 |
| Gong 2016 | 1.192 | 1.087 | 1.307 | 3.736 | 0.000 |
| Tabassum 2016 | 1.201 | 1.096 | 1.317 | 3.923 | 0.000 |
| Bronsveld 2017 | 1.196 | 1.093 | 1.309 | 3.893 | 0.000 |
| Crispo 2017 | 1.207 | 1.103 | 1.320 | 4.107 | 0.000 |
| Maskarinec 2017 | 1.203 | 1.100 | 1.315 | 4.065 | 0.000 |
| Palmer 2017 | 1.203 | 1.101 | 1.313 | 4.107 | 0.000 |
| Fang 2018 | 1.201 | 1.101 | 1.309 | 4.130 | 0.000 |
| He 2018 | 1.202 | 1.103 | 1.310 | 4.189 | 0.000 |
| Pan 2018 | 1.202 | 1.104 | 1.309 | 4.237 | 0.000 |
| Maskarinec Ice 2019 | 1.199 | 1.102 | 1.304 | 4.210 | 0.000 |
| Maskarinec Ja 2019 | 1.196 | 1.101 | 1.299 | 4.224 | 0.000 |
| Rastad 2019 | 1.193 | 1.098 | 1.295 | 4.184 | 0.000 |
| Sanderson 2019 | 1.187 | 1.094 | 1.289 | 4.105 | 0.000 |
| Vatseba 2019 | 1.209 | 1.114 | 1.312 | 4.539 | 0.000 |
| Bjornsdottir 2020 | 1.204 | 1.112 | 1.304 | 4.583 | 0.000 |
| Rennert 2020 | 1.200 | 1.109 | 1.298 | 4.531 | 0.000 |
| Park 2021 | 1.195 | 1.106 | 1.292 | 4.489 | 0.000 |
| | 1.195 | 1.106 | 1.292 | 4.489 | 0.000 |

Cumulative risk ratio (95% CI)

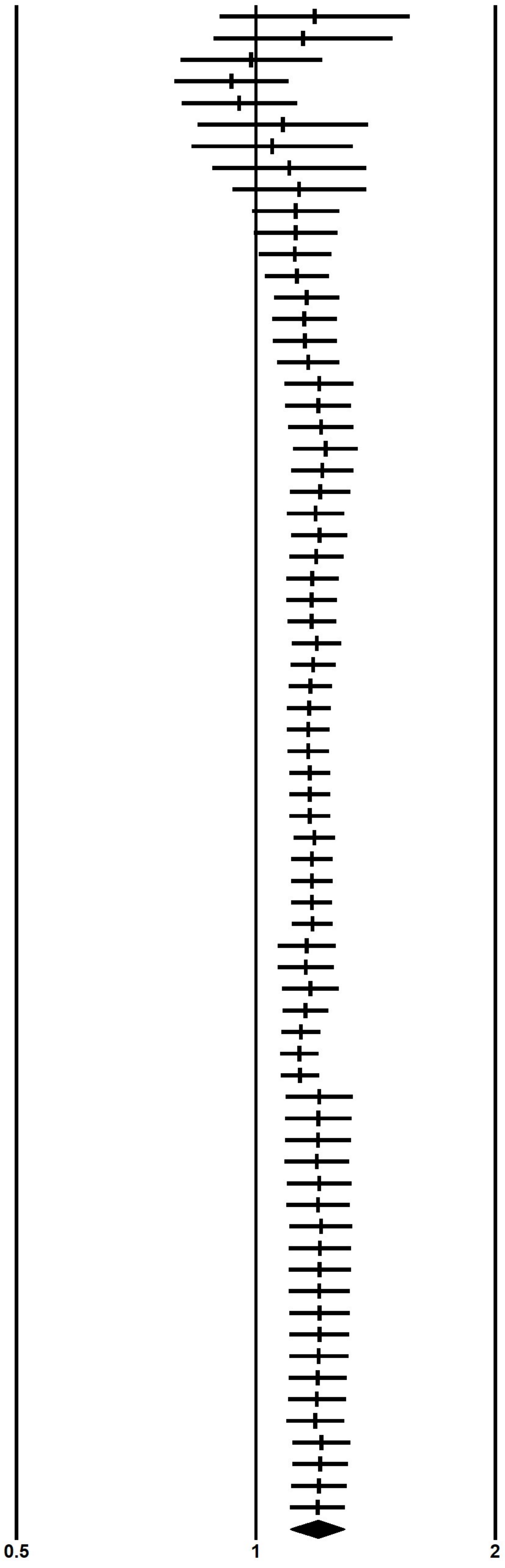
