## Supplementary figures and images for "Diabetes and Incidence of Breast Cancer and its Molecular Subtypes: A Systematic Review and Meta-Analysis"

### Supplementary Figure 3

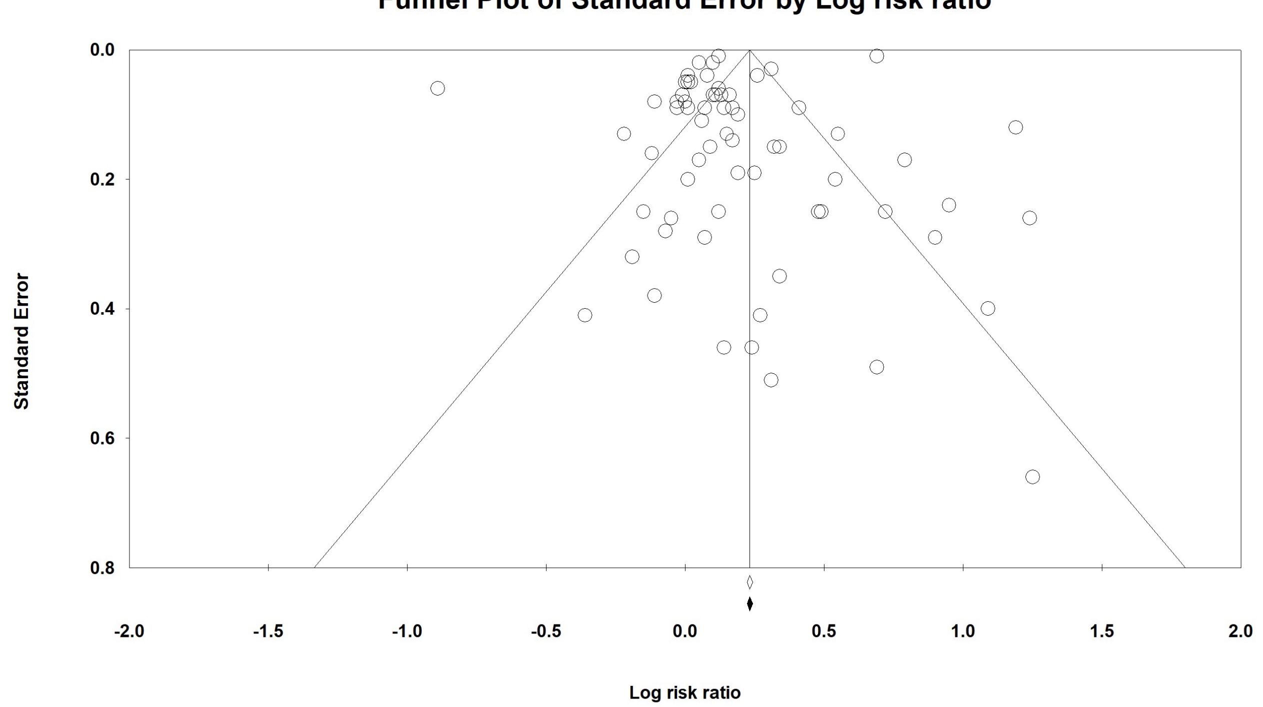

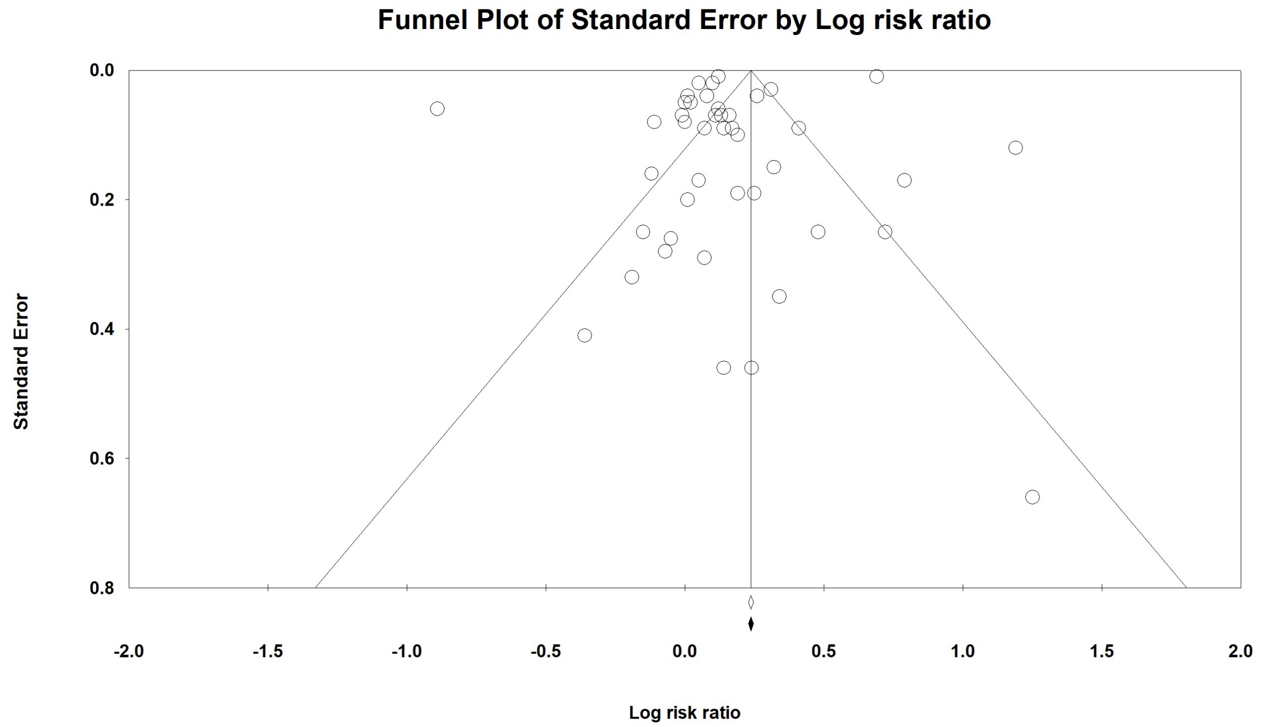

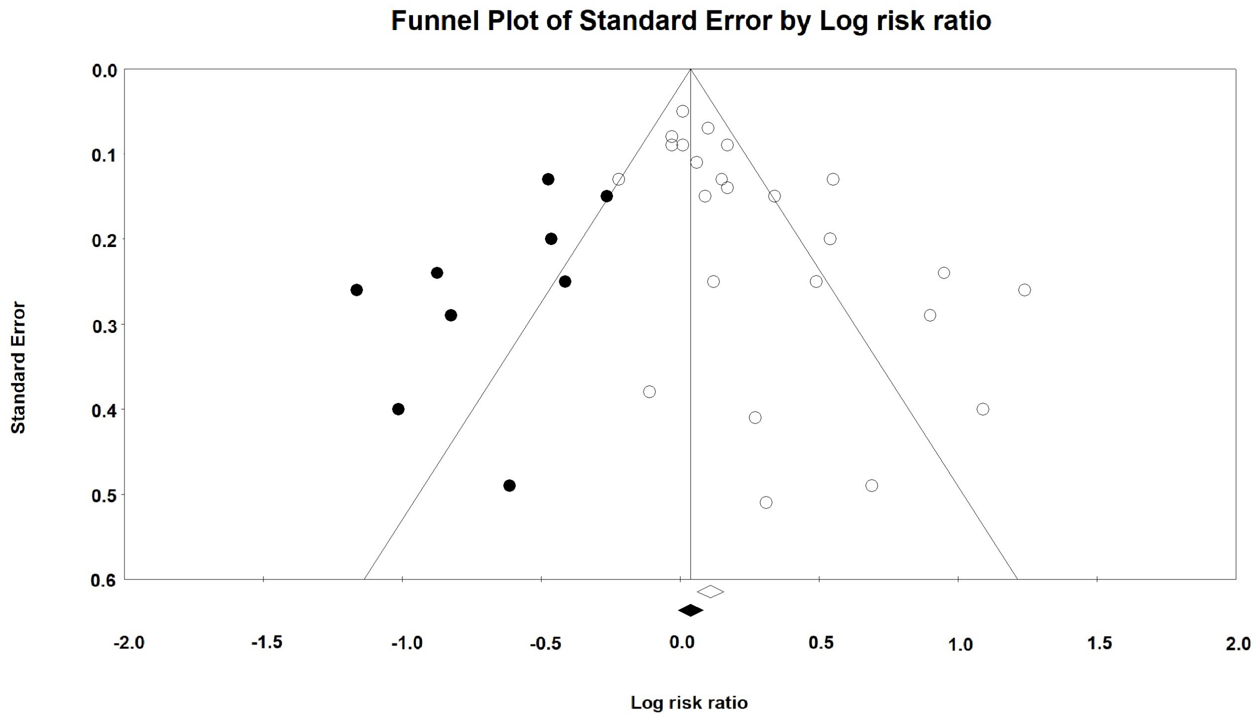
