## Supplementary material for "Diabetes and Incidence of Breast Cancer and its Molecular Subtypes: A Systematic Review and Meta-Analysis": Search appendix

| DATABASE | SEARCH STRATEGY |
| --- | --- |
| PubMed | ("Diabetes Mellitus"[Mesh] OR “type 2 diabetes”[tiab] OR “type 1 diabetes”[tiab]) AND ("Breast Neoplasms"[Mesh] OR “breast cancer”[tiab] OR “breast neoplasms”[tiab] OR "Triple Negative Breast Neoplasms"[tiab] OR “triple negative breast cancer”[tiab]) AND ("Risk"[Mesh] OR risk[tiab] OR "incidence"[mh] OR incidence[tiab] OR "Odds Ratio"[Mesh] OR odds[tiab] OR “hazard ratio”[tiab] OR “standard incidence ratio”[tiab] OR "Association"[Mesh] OR association[tiab] OR correlation[tiab]) |
| Embase | ('diabetes mellitus'/exp OR 'diabetes mellitus' OR 'non insulin dependent diabetes mellitus'/exp OR 'non insulin dependent diabetes mellitus' OR 'insulin dependent diabetes mellitus'/exp OR 'insulin dependent diabetes mellitus') AND ('breast cancer'/exp OR 'breast cancer' OR 'triple negative breast cancer'/exp OR 'triple negative breast cancer') AND ('risk'/exp OR 'risk' OR 'incidence'/exp OR 'incidence' OR 'odds ratio'/exp OR 'odds ratio' OR 'hazard ratio'/exp OR 'hazard ratio' OR 'standard incidence ratio' OR 'association'/exp OR 'association' OR 'correlation'/exp OR 'correlation') |
| Web of Science | ("Diabetes Mellitus" OR “type 2 diabetes” OR “type 1 diabetes”) AND (“breast cancer” OR “breast neoplasms” OR "Triple Negative Breast Neoplasms" OR “triple negative breast cancer”) AND (risk OR incidence OR odds OR “hazard ratio” OR “standard incidence ratio” OR association OR correlation) |
